# Genetic disease risks of under-represented founder populations in New York City

**DOI:** 10.1101/2024.09.27.24314513

**Authors:** Mariko Isshiki, Anthony Griffen, Paul Meissner, Paulette Spencer, Michael D. Cabana, Susan D. Klugman, Mirtha Colón, Zoya Maksumova, Shakira Suglia, Carmen Isasi, John M. Greally, Srilakshmi M. Raj

## Abstract

The detection of founder pathogenic variants, those observed in high frequency only in a group of individuals with increased inter-relatedness, can help improve delivery of health care for that community. We identified 16 groups with shared ancestry, based on genomic segments that are shared through identity by descent (IBD), in New York City using the genomic data of 25,366 residents from the All Of Us Research Program and the Mount Sinai Bio*Me* biobank. From these groups we defined 8 as founder populations, mostly communities currently under-represented in medical genomics research, such as Puerto Rican, Garifuna and Filipino/Pacific Islanders. The enrichment analysis of ClinVar pathogenic or likely pathogenic (P/LP) variants in each group identified 202 of these damaging variants across the 8 founder populations. We confirmed disease-causing variants previously reported to occur at increased frequencies in Ashkenazi Jewish and Puerto Rican genetic ancestry groups, but most of the damaging variants identified have not been previously associated with any such founder populations, and most of these founder populations have not been described to have increased prevalence of the associated rare disease. Twenty-five of 51 variants meeting Tier 2 clinical screening criteria (1/100 carrier frequency within these founder groups) have never previously been reported. We show how population structure studies can provide insights into rare diseases disproportionately affecting under-represented founder populations, delivering a health care benefit but also a potential source of stigmatization of these communities, who should be part of the decision-making about implementation into health care delivery.

**Author Summary:** It is well recognized that genomic studies have been biased towards individuals of European ancestry, and that obtaining medical insights for populations under-represented in medical genomics is crucial to achieve health equity. Here, we use genomic information to identify networks of individuals in New York City who are distinctively related to each other, allowing us to define populations with common genetic ancestry based on genetic similarities rather than by self-reported race or ethnicity. In our study of >25,000 New Yorkers, we identified eight highly-interrelated founder populations, with 202 likely disease-causing variants with increased frequencies in specific founder populations. Many of these population-specific variants are new discoveries, despite their high frequency in founder populations. Studying recent genetic ancestry can help reveal population-specific disease insights that can help with early diagnosis, carrier screening, and opportunities for targeted therapies that all help to reduce health disparities in genomic medicine.

## Introduction

Rare diseases collectively occur in 3.5-5.9% of the population(1) They involve significant morbidity and mortality, risk to family members and socio-economic consequences, and thus have the characteristics typical of a public health priority. Responding to this public health issue by studying rare diseases on a population scale is challenging because of the difficulty identifying individuals and families with uncommon conditions that are often refractory to diagnosis. An eventual solution will involve widespread application of sequencing of patients’ entire genomes in health care with sensitive and high-confidence prediction of damaging DNA sequence variants, but this remains a remote goal at present. In the interim, a typical approach in clinical practice is to use a person’s ‘genetic ancestry group’ (2) to highlight the rare diseases that are more common in that community and could be affecting the patient presenting for care. Populations that experienced small population size in the past tend to have enrichment for otherwise rare genetic conditions due to a ‘founder effect’, as exemplified in Ashkenazi Jewish individuals for their well-characterized set of genetic conditions (3,4) By identifying other populations with founder effects, the genetic conditions more likely to occur in individuals from those communities can also be defined, and clinicians who serve these communities can be prepared to look out for these conditions. Extending the insights into rare disease risks for genetic ancestry groups other than White Europeans has been limited by the failure to include non-European populations in genomics research (5,6) This bias magnifies health disparities and impedes effective delivery of medical care to marginalized groups and underserved populations. Recognizing this neglect, the All of Us (AoU) Research Program in the United States has been designed to represent the country’s diversity (7,8) In this study, we focused on the genomes of individuals in New York City (NYC), representing a diverse and admixed urban population studied extensively through AoU as well as the separate Bio*Me* biobank(9,10). We show how population genetics approaches using these data resources are able to reveal previously undiscovered rare disease susceptibilities in diverse genetic ancestry groups, particularly those with a founder effect. We were able to define groups with increased ‘genetic similarity’(2) and characterize population structure by identifying segments of DNA that are shared among individuals due to inheritance from a common ancestor, also called identity-by-descent (IBD). This has previously been performed successfully in cohorts of different genetic ancestries (10–14) with implications for understanding population-specific disease risk (9,10,13) Here we explicitly test the association between population structure and disease risk by focusing on population-specific enrichment of variants curated as disease-causing in the ClinVar database (15) The results show how health systems and providers can benefit from recognizing rare diseases in the populations they serve, including the potential benefits of early detection of rare diseases as well as prenatal carrier screening in these communities, and the targeted use of specific therapies.

## Results

### The population structure of NYC participants of the All of Us Research Program

We studied the genetic diversity of NYC residents using genetic data from 13,817 participants of the AoU Research Program. This dataset excludes ‘related’ individuals, those who are second cousins or closer. We found that the AoU cohort in NYC is diverse across the five boroughs (**Fig. 1a**, **Figure S1**), and that the proportions of self-reported race/ethnicity information for each borough are comparable to those from census data (**Figure S2a**) (16). Of the boroughs, Manhattan and the Bronx are over-represented (**Figure S2b**). Admixed individuals accounted for a large proportion of the dataset (**Figure S1**).

**Figure 1:**
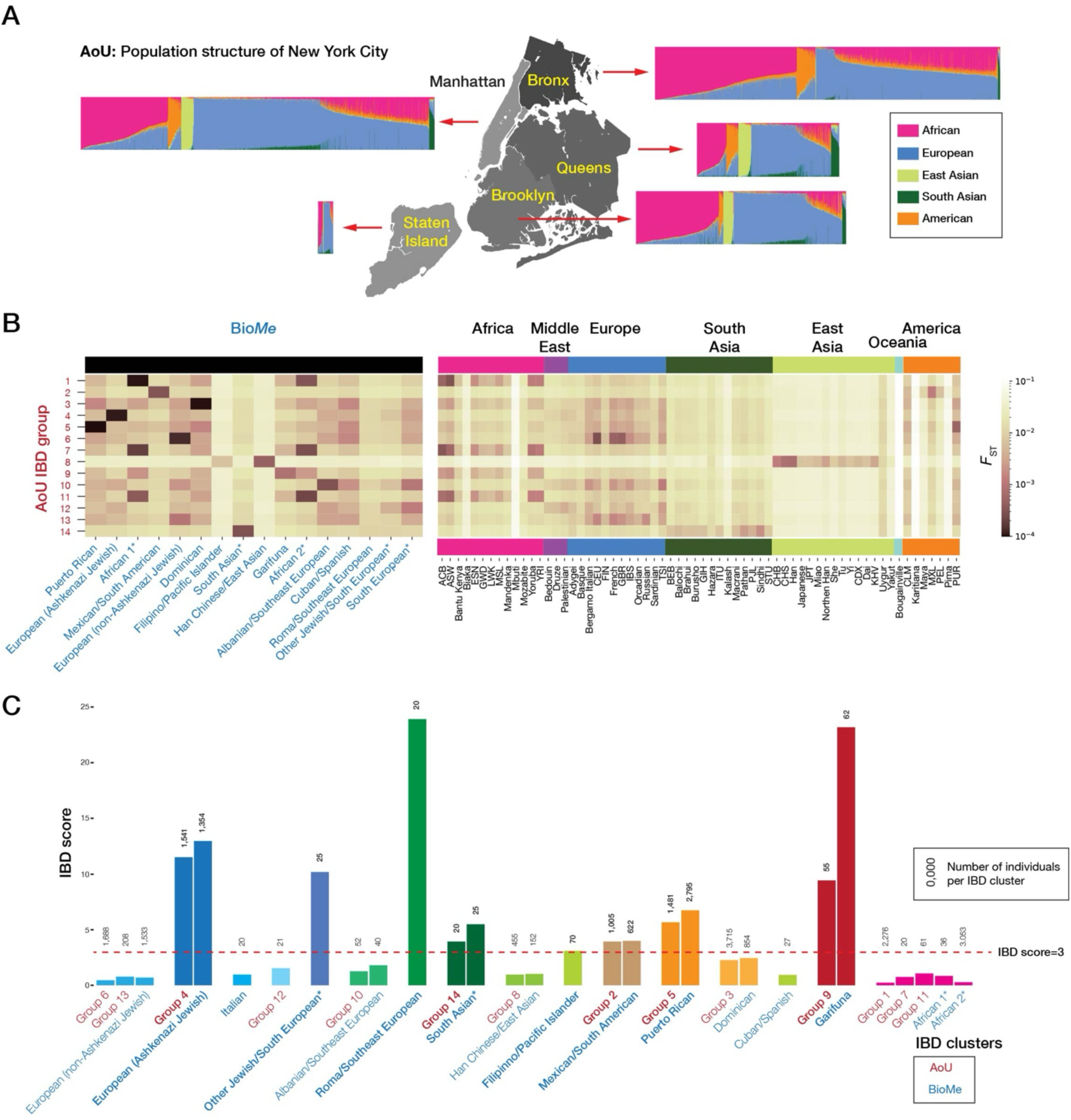
Population membership and geospatial distribution of All of Us IBD groups. **(A)** The geospatial distribution of all AoU individuals across NYC boroughs; **(B)** Pairwise *F*_ST_ comparisons between AoU IBD groups, Bio*Me* IBD groups and global reference populations; **(C)** IBD scores for AoU and BioMe IBD groups, with sample sizes above each bar. IBD group with *F*_ST_ < 0.001 are indicated by the same color, bold fonts identify founder populations. The red dotted line indicates an IBD score of 3, which is the cut-off value we used to define a founder population (**Fig. 2**); Only IBD groups with 20 and more individuals are shown to protect participants’ privacy based on the AoU Data and Statistics Dissemination Policy. An asterisk next to labels represents populations with inadequate reference information for annotation.

We constructed a network based on identity-by-descent (IBD) sharing to capture fine-scale recent population structure in the AoU NYC participants. After filtering edges to only reflect recent shared ancestry and exclude close familial ties, 98.6% of the cohort was included in the network. Among these individuals, we identified 14 IBD groups with a minimum of 20 individuals each (**Figure S1b**), representing 91% of the AoU NYC cohort. To allow comparison of our AoU results with results using the independent NYC Bio*Me* biobank, we replicated the AoU IBD analysis on Bio*Me.* The network included 95.6% of the Bio*Me* cohort. We identified 16 groups with ≥20 individuals representing 92.5% of the Bio*Me* cohort (**Fig. 1b**), consistent with their published results (9,10). We found that AoU and Bio*Me* have several similar populations with *F_ST_* < 0.001 between them, even after removing related individuals across both datasets, reflecting their shared NYC recruitment area (**Fig. 1b**).

Of the IBD groups, there were five from AoU and eight from Bio*Me* with IBD scores >3, defining them as founder populations (**Fig. 1c**). Of the eight founder groups from BioMe, founder effects in Ashkenazi Jewish, Puerto Rican and Garifuna were reported previously (9,10). We also found the founder populations in AoU showed major geospatial differences, with the IBD group 9 (Garifuna) in particular over-represented in the Bronx compared to other boroughs (not shown to comply with AoU Data and Statistics Dissemination Policy).

### Detection of founder populations in NYC

Since our AoU dataset and Bio*Me* are both NYC cohorts with shared genetic features (**Figs. 1b-c, Figure S3**), we combined the IBD groups from AoU and Bio*Me* with pairwise Hudson’s *F_ST_* values < 0.001, resulting in 16 IBD groups which we annotated based on inferred ancestry (**Fig. 2**). Eight groups were identified as founder populations (IBD score >3). The populations were named based on self-defined ancestry as provided by the reference datasets, when available (**Figs. 1b-c,2,3, Table S1**). Genetic ancestry also acts on a continuum(17), therefore some IBD groups appeared to be more discrete (*e.g.* Garifuna, Puerto Rican), whereas others include individuals across a broader geographic range (*e.g.* Filipino/Pacific Islander, Mexican/South American, Han Chinese/East Asian). In situations where groups could not be confidently labeled, the closest associated population group was used as a placeholder label until more genomic reference datasets become available. These populations are labeled with an asterisk (**Figs. 1b-c, 2,3**).

**Figure 2:**
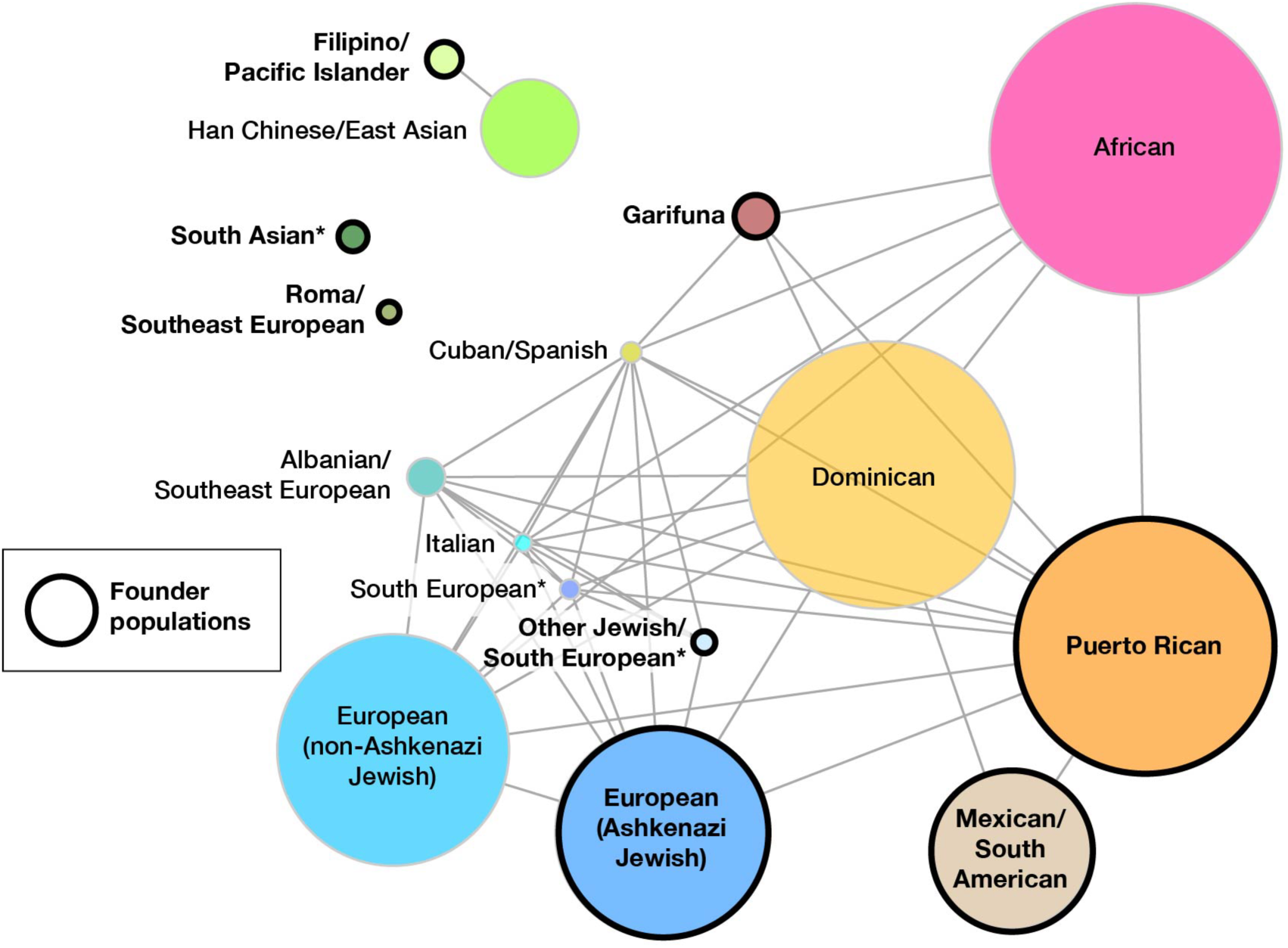
IBD groups identified in NYC from the combined AoU and Bio*Me* datasets. A network depiction of the IBD groups based on *F*_ST_ between IBD groups. Edges were weighted by logarithm of *F*_ST_. Only edges representing *F*_ST_ <0.01 are shown, with founder populations circled in black. Circle sizes reflect the number of individuals in each IBD group. An asterisk next to labels represent populations with inadequate reference information for annotation.

### Identification of pathogenic founder variants

We then studied the genomes of the members of the founder populations to identify pathogenic variants characterizing each group. We used the ClinVar resource (15) as a curated source of disease-causing variants, while recognizing that the bias of ClinVar towards documentation of variants in individuals of European ancestry (18). We focused on recurrent, rare, disease-causing variants, given our focus on founder effects. By requiring the pathogenic/likely pathogenic (P/LP) variant to occur in at least 2 individuals not from the same family, we identified 3,616 P/LP recurrent variants in NYC individuals. Consistent with the known ClinVar bias(18), European ancestry IBD groups showed more pathogenic variants than other IBD groups, especially when compared with individuals with African ancestry (**Fig. 3**).

**Figure 3:**
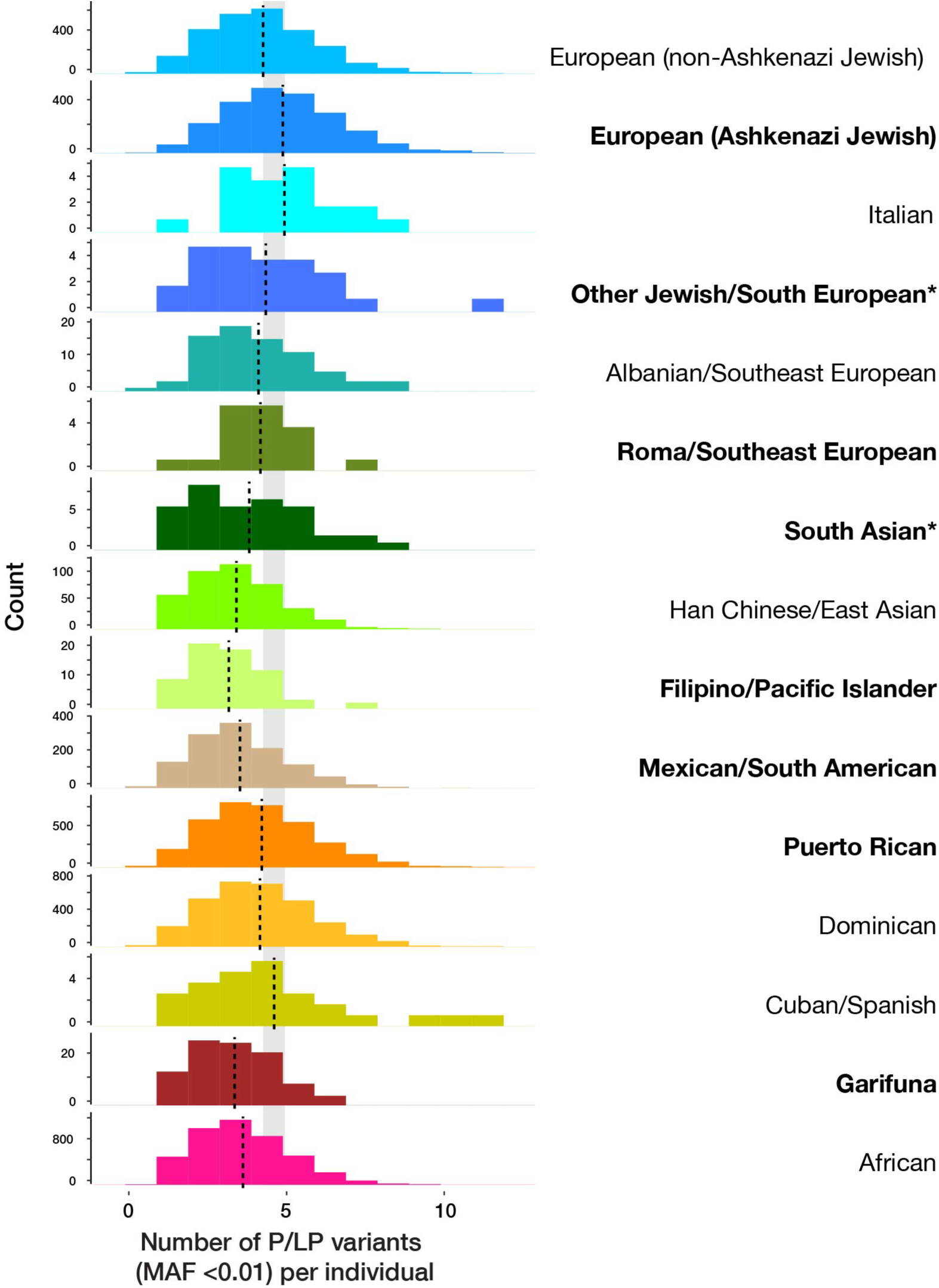
Number of P/LP variants per individual for each NYC IBD group identified from the AoU and Bio*Me* datasets. Each histogram shows the number of P/LP variants with minor allele frequency < 0.01 per individual for 15 IBD groups. The vertical dashed line indicates the mean value in each group. The ‘South European’ IBD group only included WGS data for fewer than 20 individuals and was removed due to the All of Us Data and Statistics Dissemination Policy. The shaded gray rectangle represents the range of mean values for the uppermost three European groups, highlighting the lower number of ClinVar variants annotated in those of Asian, American and African ancestries. Asterisk next to labels represent populations with inadequate reference information for annotation.

We detected 674 unique P/LP variants significantly enriched across the 8 founder populations (Fisher’s Exact *p* > 0.05) (**Table S2**) with 202 of these variants passing Bonferroni correction. Of the 674 variants, 478 variants have two or more ClinVar gold stars, meaning variants are from practice guidelines, reviewed by expert panel, or from multiple submitters with evidence and no classification conflicts. **Table S**Those variants with no ClinVar gold stars should in general be interpreted with caution, such as the *KRT18* variant not previously described to be common in Ashkenazi Jewish individuals. The *CD55* variant associated with protein-losing enteropathy (19) and shown in cell studies to cause loss of CD55 on the cell surface (20) also lacks a star rating, illustrating how the absence of this rating should not be used to exclude variants as disease-causing. This *CD55* variant also does not pass Bonferroni correction, nor do known founder effect variants in *HBB* in the South Asian group and a *SLC26A4* variant in the Filipino/Pacific Islander group, prompting us to include variants that do not pass multiple testing correction in **Table 1** as candidate founder disease-causing variants in the IBD groups. We identified 51 variants from this broader list that have minor allele frequencies of > 0.005 in one or more IBD groups, the Tier 2 threshold for inclusion into prenatal screening panels (21). Of these, 25 are new, previously unrecognized founder effect variants (**Table 1).** The results shown in **Fig. 3** support the likelihood that the numbers of P/LP variants in non-European groups are likely to represent an underestimate, and that more disease-causing variants remain to be discovered in these under-studied groups.

**Table 1:**
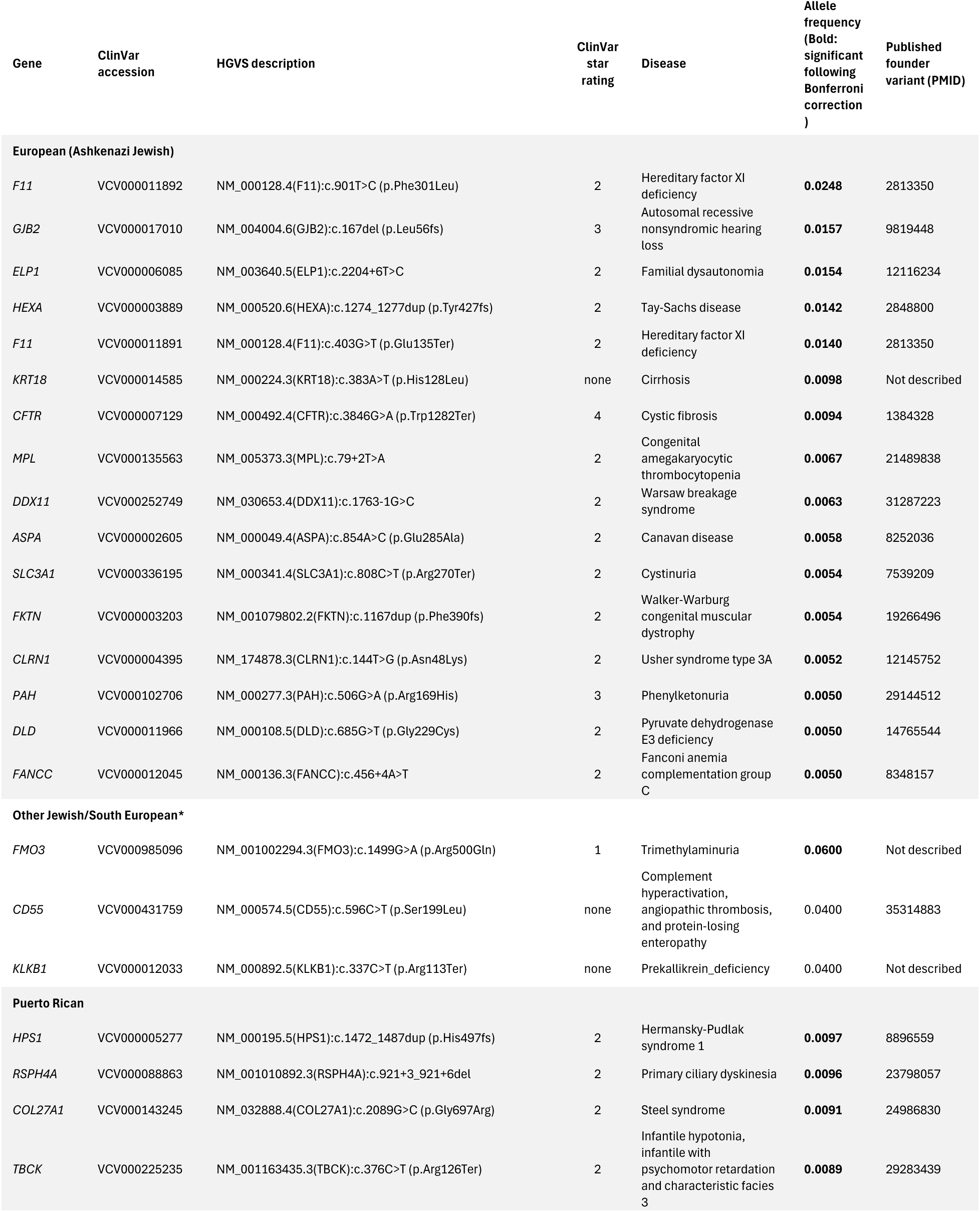
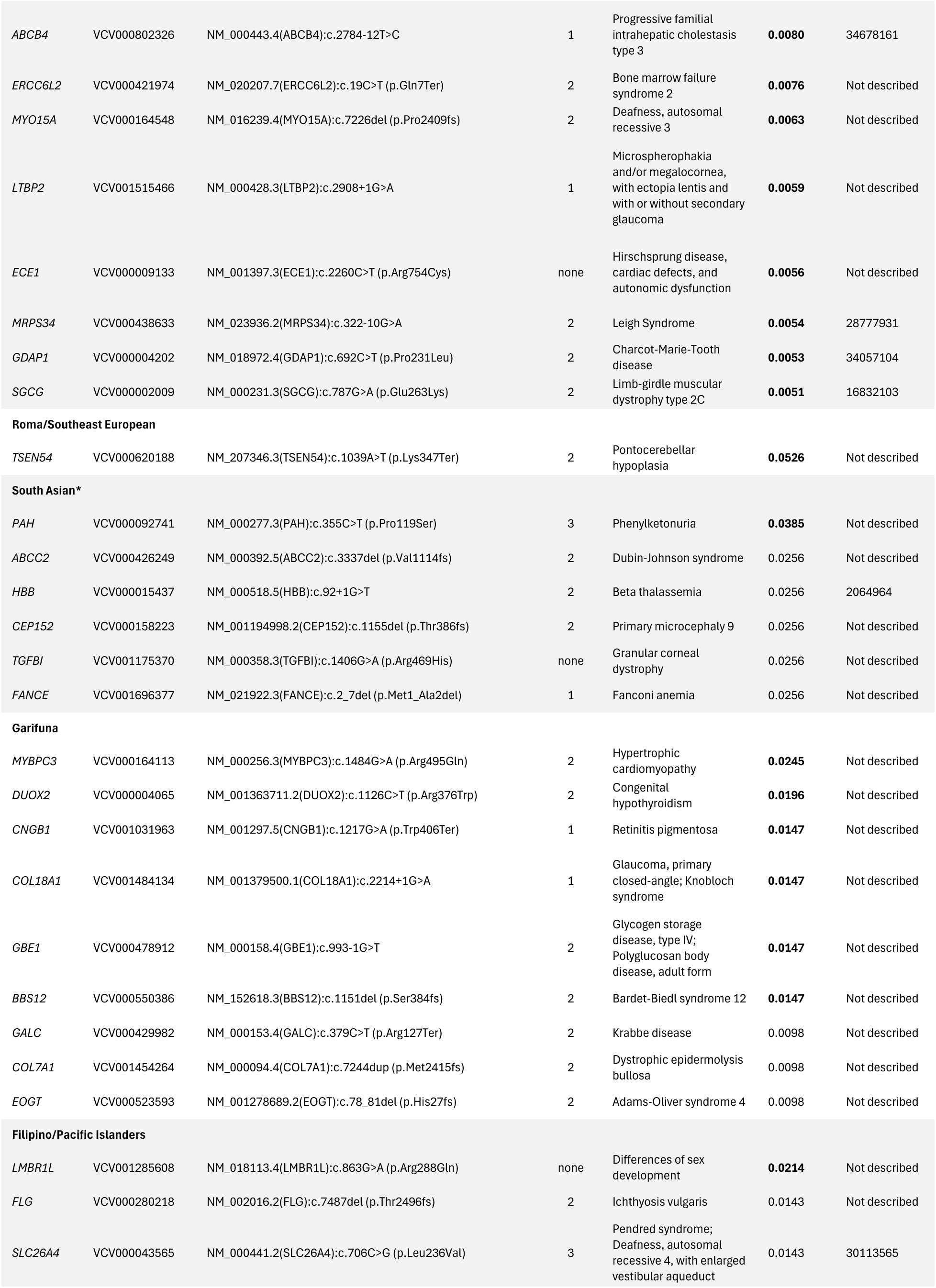
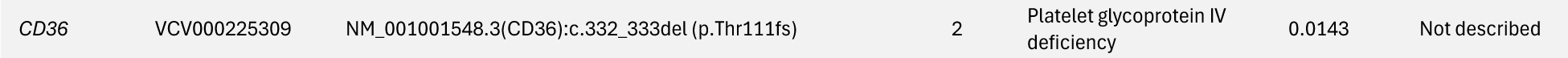
ClinVar P/LP variants with allele frequencies exceeding 1/200 within seven founder populations in NYC. The table shows the P/LP variants that occur within each population at a frequency of at least 1/200 alleles, the threshold for inclusion in prenatal testing. Those with allele frequencies in bold type pass Bonferroni correction. Of these 51 variants, 26 have already been published as founder mutations, especially in the European (Ashkenazi Jewish) and Puerto Rican populations. The other 25 are new, previously unrecognized founder effect variants. Abbreviations: PMID, PubMed reference number. An asterisk next to labels represent populations with inadequate reference information for annotation. Some of the allele frequency counts displayed here represent <20 AoU participants. The AoU Resource Access Board reviewed this work and granted an exception to their Data and Statistics Dissemination policy to report these frequencies.

### Ancestry analysis for shared founder variants in individuals of Caribbean ancestry

We detected 12 and 9 founder P/LP variants for Puerto Rican and Garifuna IBD groups, including variants that did not pass Bonferroni correction, respectively (**Table 1**). Of these 21 variants, 15 were also detected at lower frequencies in other IBD groups **(Table S3)**. To test whether this was due to shared ancestry, we inferred local ancestry (the origin of the DNA containing each P/LP variant) in the Caribbean individuals to identify the ancestral population in which each P/LP variant arose originally. We found the majority of the founder variants in Puerto Ricans to be located on haplotypes of European ancestry, with the remaining founder variants located on African and Indigenous American haplotypes (**Table S3**). Of these, the origin of the *COL27A1* Steel syndrome variant on chromosome 9 has also previously been characterized as Native American ancestry (22) We illustrate the sharing of founder effect mutations across Caribbean groups and with European and African groups in **Fig. 4**. The *MYBPC3* variant that occurs frequently in the Garifuna was in an African haplotype, but was also found in a European haplotype in an individual unassigned to any IBD group, indicating that the same mutation arose independently in African and European individuals. This finding demonstrates how local ancestry analysis can be used to reveal the evolutionary history of founder effect mutations, and how the presence of a founder effect mutation does not by itself indicate a person is part of a known founder population.

**Figure 4:**
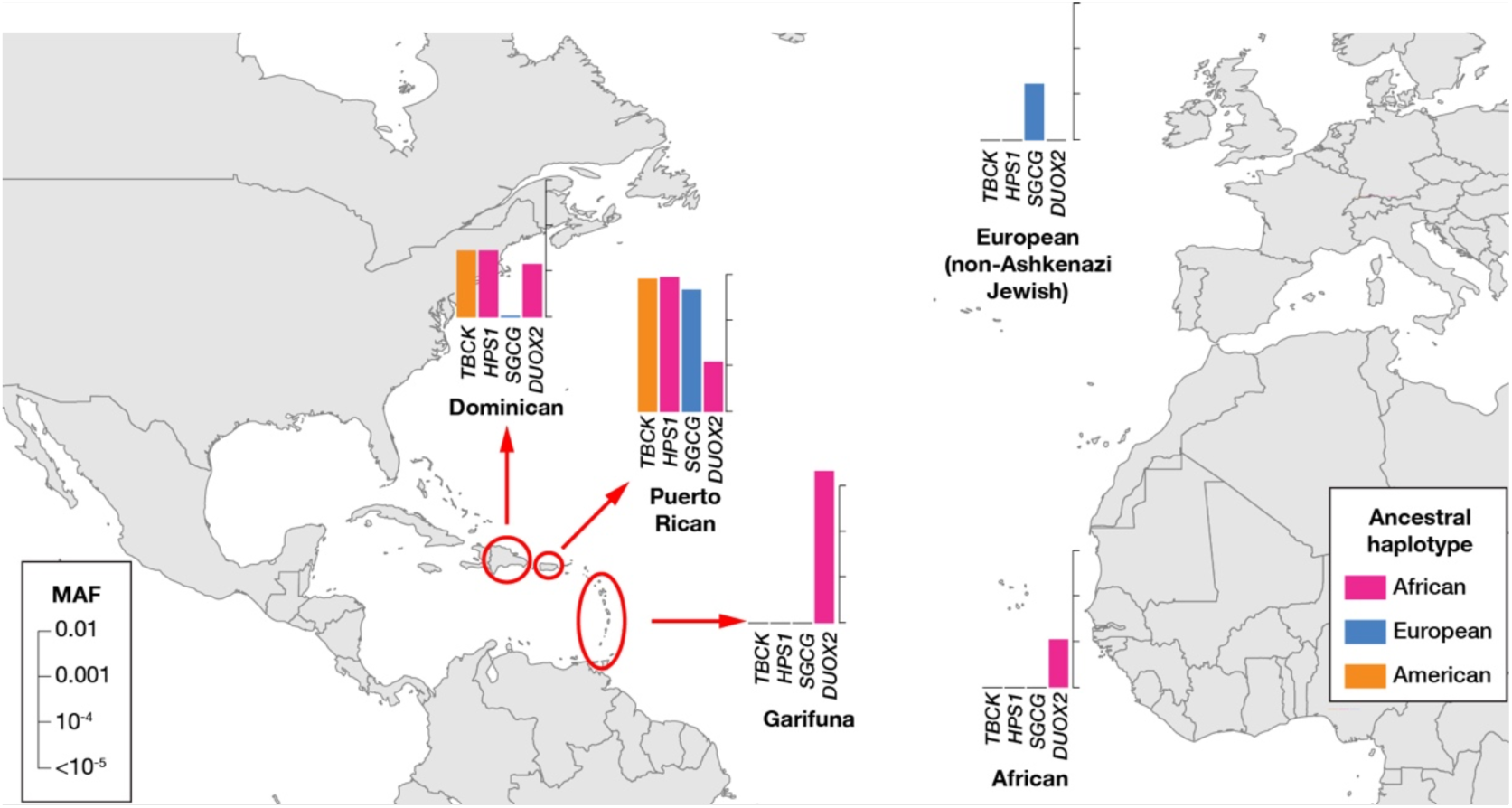
Shared founder variant frequencies in Caribbean IBD groups. Four examples of founder mutations are illustrated with comparisons of frequencies in other IBD groups. Local ancestry analysis reveals African origin of the *HPS1* and *DUOX2* pathogenic variants, the Indigenous American origin of the *TBCK* and European origin of the *SGCG* pathogenic variants. No variant is exclusive to one IBD group but occurs across multiple Caribbean groups, reflecting the complex ancestries of these populations and the weakness of demographic categories such as race and ethnic origin as the sole predictors of genetic disease risks.

## Discussion

In this study, we showed that founder populations exist in the megacity of New York, and that the individuals from these genetic ancestry groups have distinctive increased risks of rare genetic diseases. The deliberate inclusiveness of the AoU Research Program(23) has ensured that insights extend to communities with genetic ancestries other than those included in the non-Hispanic White demographic. By defining genetic similarity using IBD sharing network as opposed to crude demographic or continental groupings, and focusing on DNA sequence variants with strong prior evidence for causing genetic diseases, we rediscovered many known rare disease-causing variants common in the better-studied Ashkenazi Jewish and Puerto Rican communities, while revealing new founder mutations in these and other founder populations in NYC. The value of this recognition is not only in terms of otherwise rare diseases entering into the differential diagnosis of a patient being evaluated clinically, but also in terms of inclusion of these genes and conditions in prenatal carrier screening. The utility of information about rare genetic conditions in a founder population is exemplified in the Ashkenazi Jewish community, where genetic testing panels for prenatal use is expanding to include presymptomatic testing for conditions affecting the parents (24) The American College of Medical Genetics has recommended that “carrier screening paradigms should be ethnic and population neutral and more inclusive of diverse populations to promote equity and inclusion” (21) This study demonstrates how current carrier screening panels can be expanded to serve a broader set of under-represented communities, using the example of the diverse population of NYC.

To identify genetic similarities between individuals, we used an IBD sharing network, which can identify groups of individuals who share recent ancestry in an unbiased manner (10–14) Due to the nature of IBD segments, this approach works well to identify founder populations, who are expected to share higher genetic ancestry as well as population-specific disease-causing variants. However, the assignment of individuals to a group within the network is highly dependent on who is included in the network. This becomes more complicated when there are individuals who lie at the boundaries between groups, because they have multiple ancestry components due to admixture. The inclusion of admixed individuals with shared ancestry allows us to capture more population-specific pathogenic variants but also leads to underestimations of allele frequencies. Varying the length thresholds of IBD segments, using different community detection algorithms, or combining or annotating IBD groups based on different *F*_ST_ thresholds will also change the resolution of population structure that is captured.

To describe these groups, we used information about how the individuals from these groups described themselves as well as genetic similarity with reference population (*F*_ST_), defining the genetic ancestry groups for this study (**Table S1)**. Prior reports of disease-causing DNA sequence variants allowed inference of the origins of some of the founder populations, with the *SLC26A4* variant in the Filipino/Pacific Islander group known to be common in Filipinos (25), and the *CD55* variant in the Other Jewish/South European group previously identified in Bukharian Jewish individuals (19) While genetic ancestry group information is not routinely captured in health records, a clinical encounter seeking to understand a patient’s rare disease will typically involve a detailed family history, seeking evidence of consanguinity that involves asking about the origins of grandparents and prior generations. Genetic ancestry group information is therefore much more likely to be captured in rare disease care and can be used to raise the possibility that known founder mutations in that community may be the cause of the affected individual’s rare disease.

The potential that the recognition of the presence of a rare disease in a founder population can lead to targeted therapies is exemplified by the *CD55* variant in the Bukharian Jewish community, which can cause a spectrum of presentations from mild abdominal discomfort following a high-fat meal to a severe syndrome including protein-losing enteropathy, and is effectively treated by the complement C5-inhibitor eculizumab (26) Implementation in health care systems of information about rare disease susceptibility for founder populations can therefore encompass prenatal screening, clinical decision support to prompt clinicians to be aware of an otherwise rare disease in a patient from a defined community, and can lead to therapeutic interventions.

For a variant to be categorized in ClinVar as Pathogenic or Likely Pathogenic, it has to fulfil a number of stringent criteria (27) The degrees of confidence about the variant categorization is represented by a star system, reflecting the degrees of expert curation of the variant. We note that the *KRT18* variant that meets the criteria for inclusion in **Table 1** is rated with zero stars, and may not be a true risk allele in the European (Ashkenazi Jewish) genetic ancestry group. There are other reasons why categorizations of likely pathogenic or pathogenic variants in ClinVar (15) may not be reliable. For example, older submissions to the database are prone to subsequent conflicting interpretations (28) sometimes because of the failure to appreciate the variant to be relatively common in one understudied population at the time of submission (29). Our use of ClinVar has revealed many new variants causing rare genetic diseases in under-represented populations of NYC, but we recognize that ClinVar’s bias towards P/LP variants in Europeans implies that there remains even more to be discovered about rare diseases in the non-European founder populations studied.

Another general problem with pathogenicity classifications is the assumption that the presence of a variant at a frequency higher than the prevalence of the associated disease should lead to the variant being reclassified as non-causative. This by itself is a reasonable general assumption, but in the context of the groups we are studying here we find two reasons for concern. One is that founder populations can have high frequencies of a variant and should be excluded from this filtering approach (30) This approach is implemented in Grpmax FAF, the filtering allele frequencies offered by gnomAD (31) which excludes founder populations like the Amish, Ashkenazi Jewish and Finnish from frequency calculations. By identifying other founder populations, approaches like Grpmax FAF can be refined with variant frequency information from these additional groups. The other concern is that disease prevalence measurements may vary between communities depending on access to care, which is a concern in NYC (32) and in US health care more generally (33) If a community lacks access to care, the prevalence of a rare disease in that community may go unrecognized, with the further failure to recognize an association with a disease-causing variant that may then be misclassified as benign. As we demonstrate here, large-scale population studies like AoU will make it increasingly feasible to gain insights into variant frequencies in different genetic ancestry groups, including founder populations, but some of these genetic ancestry groups will also be defined by limited access to health care. There is the potential to worsen health equity by applying exclusive variant frequency thresholds that fail to recognize the genetic disease burden of founder populations through adequate phenotyping.

We have to balance the value of identifying a genetic disease risk in a community with the risk of stigmatizing that group. The AoU Research Program notes this potential for biased interpretation promoting negative stereotypes (34) We therefore emphasize how pathogenic variants occur in everyone, regardless of demographic categorization or genetic ancestry (**Fig. 3**). What distinguishes founder effect groups is not likely to be the overall burden of genetic damage, but instead the over-representation of specific genetic diseases (**Table 1**) within that burden of damaging variants. We also stress how differences in the numbers of damaging variants in the genomes of people from different parts of the world have more to do with incompleteness of information about and genomic annotations of damage. We demonstrate the shortcomings of crude demographic categories such as race and ethnic origin in predicting genetic disease risks. We identified a strong founder effect in the Garifuna ancestry group, but when they self-identify their race and ethnicity they include African American/Black, Hispanic and Latino, and in some cases diverse countries of origin, illustrating the weakness of these categorizations as proxies for genetic variation (17,35) Similarly, we found multiple self-identified ancestries in some of the non-founder groups, who were not the focus of this manuscript. For example, the ‘African’ group included individuals who align with African, African-American, and African-Caribbean ancestry groups, reflected in the wide spectrum of ancestry in the PCA analysis (Figure 1b, Figure S1b).

We find that some of the risk alleles from the founder Caribbean populations in NYC also exist at lower frequencies in other Caribbean New Yorkers, because of the complex history of pre-colonial civilization, colonization, slavery and migration. In some cases, individuals of non-Caribbean origin appeared to have founder effect variants that appear in the Caribbean, but these variants were mostly located on shared haplotypes derived from the same continental ancestry, with the exception of one variant that appears to have independently arisen in Garifuna and European individuals (**Table S3**). Demography is therefore only modestly informative in predicting disease risk, making any associated stigma tenuous. Instead we followed the guidelines of the National Academies of Sciences, Engineering, and Medicine (NASEM) on the Use of Race, Ethnicity, and Ancestry as Population Descriptors in Genomics Research (2) to quantify objectively ‘genetic similarity’ using IBD sharing, and ‘genetic ancestry’ labels using those provided by members of each group **(Table S1**). We also worked with members of the genetic ancestry groups highlighted in the results to discuss and prepare this report, following recommendation 5 of the NASEM report. These best practice guidelines are clearly of value in using population descriptors in ways that enhance the application of genomic insights in medical care delivery.

This study shows how genetic variants that cause diseases that are rare globally can be common locally within a population, and can influence the spectrum of diseases of patients served by individual health systems. Our focus was on NYC, but the same approaches can be extended nationally using AoU data and comparable international data resources. The insights gained are essential for better health care provision, while highlighting the need to gain insights into the phenotypic manifestations of disease-causing variants in marginalized populations with less access to health care.

## Materials and Methods

### Research participants and dataset preparation

Participants living in NYC in the AoU Program version 6 curated data repository (8) were identified by the first three digits of their Zip Code, allowing borough-level resolution of geographic residence. Microarray genotype data were used to assess the population structure of NYC participants by principal component analysis (PCA), by global ancestry analysis as performed by SCOPE (36) and by Identity-by-descent (IBD) analysis as described below. Samples were QCed by call rate and kinship coefficient using PLINK v2.00a2.3LM (37) No individuals were filtered out by the call rate threshold of 0.9 (—mind 0.1). To remove close relatives, either of the pairs of individuals who showed king kinship coefficients > 0.125 were removed using –king-cutoff 0.125 in PLINK2.0 (37). Variants of the array data were filtered with the following conditions using PLINK2.0: minor allele frequency >0.01, genotyping rate per site > 0.95, and p-value for the departures from Hardy Weinberg Equilibrium (HWE) > 1 × 10^−6^ (–maf 0.01 –geno 0.05 –snps-only –hwe 1e-06). After QC steps, 13,817 participants and 720,630 SNPs remained for downstream analysis.

Out of these individuals, 10,381 individuals had whole genome sequence (WGS) data available. We used this subset of individuals and whole genome sequence data from an independent NYC biobank, the Mount Sinai Bio*Me* biobank (9,10), to identify founder pathogenic variants. Approval to study these de-identified data was granted by the Albert Einstein College of Medicine Internal Review Board (Protocol 2016-7099). All analyses on AoU participants included in the manuscript were also approved by the All of Us Resource Access Board.

### Comparison of demography between US Census and AoU NYC participants

To show the extent to which our dataset represents the demography of NYC, we compared the proportion of four major self-described race and ethnicities per borough between census data and AoU NYC participants. We obtained census data from the following source: https://www.census.gov/quickfacts/fact/table/richmondcountynewyork,newyorkcountynewyork,q ueenscountynewyork,kingscountynewyork,bronxcountynewyork,newyorkcitynewyork/PST04522 (16).

For AoU NYC participants, self-identified race/ethnicity was obtained using a questionnaire. Participants answered the question: “Which categories describe you? Select all that apply. Note, you may select more than one group.” in the Basics Survey. Borough residence was defined based on the first three digits of the zip code of residence, provided by AoU.

### PCA and global ancestry analysis

PCA and global ancestry analysis were conducted using PLINK 2.0 and SCOPE (36) in supervised mode, respectively, on a combined dataset comprising 13,817 AoU participants and 3,584 individuals from the 1000 Genomes Project (1KGP) (38) the Human Genome Diversity Project (HGDP) (39) and the Simons Genome Diversity Project (SGDP) (40) using a total of 150,213 SNPs. Prior to global ancestry analysis using SCOPE, we conducted ADMIXTURE analysis (41) with K=5 on this assembled reference panel, and further identified individuals within this panel for whom >95% of their genomes appeared to originate in any of five continental ancestries: African, European, South Asian, East Asian and Native American (38) The supervised SCOPE analysis was run based on this subset of reference panel participants.

### Identity-by-descent (IBD) analysis

IBD groups, the sets of individuals who share ancestry as defined by shared IBD segments, were constructed from the microarray genotypes. Phasing of the genotypes was conducted with Beagle v5.4 (42) using all populations from 1KGP (38) as references. We used Templated Positional Burrows-Wheeler Transform (TPBWT) (43) on the phased dataset to infer IBD segments >3 cM across all pairs of individuals. The total length of IBD sharing for all pairs of individuals was used to construct an undirected network using the iGraph package(44) in R. To focus on recent demography and to reduce clustering of extended families, we filtered for edges with cumulative IBD sharing ≥12 cM and ≤72 cM, as previously described (11,14) IBD groups were detected using the infomap.community() (45) function on the constructed network using default parameters. To assess the strength of the founder effect for each IBD group, we estimated the ‘IBD score’, the average length of IBD segments between 3–20 centimorgans (cM) shared between two genomes normalized to sample size, as previously described (46) We also estimated the IBD score per group and per borough. To confirm the robustness of our approach and to obtain a reliable reference for population labels, we conducted IBD sharing network analysis for an independent NYC cohort, the Mount Sinai BioMe Biobank9(9,10) (dbGaP Accession number phs001644). After QC, the BioMe dataset consisted of 11,549 individuals and 982,770 SNPs. We performed IBD analysis as above and named each BioMe IBD group based on individuals‘ detailed self-reported ethnicity provided separately by BioMe leadership (Alexander Charney, personal communication). IBD groups in BioMe and AoU with IBD score >3 were defined as founder populations (**Fig. 1c**).

### Inferring ancestral background of individuals in IBD groups

Since AoU did not provide the detailed ethnicity information that is particularly useful for defining founder groups, we inferred population ancestry (*e.g.* Puerto Rican, Dominican, Ashkenazi Jewish) in AoU IBD groups by estimating Hudson’s *F*_ST_ between each group and populations in genomic reference panels using PLINK2.0. The reference panel included 14,985 individuals and 140,952 biallelic SNPs from global populations with sample sizes > 10 individuals in 1KGP, HGDP and SGDP together with IBD groups in Bio*Me*. We also conducted PCA for the merged dataset. IBD groups in AoU and Bio*Me* with *F*_ST_ values < 0.001 were combined in further analyses.

### Detection of pathogenic founder variants for rare diseases

We extracted variants categorized as pathogenic or likely pathogenic (P/LP) in the ClinVar database (15) (version ClinVarFullRelease_2023-01.xml) from WGS data of 10,381 NYC AoU participants and 11,549 Bio*Me* participants. Of the 193,935 P/LP variants registered in ClinVar as of Jan 7, 2023, we detected 27,125 variants in our NYC cohort. We removed close relatives and excluded variants that appeared only once in the NYC dataset. We then filtered variants with genotype rate < 0.9 and p-values for departures from Hardy-Weinberg Equilibrium (HWE) <1 × 10^−16^. We set a small HWE threshold anticipating that rare variants may likely diverge from HWE due to high heterozygosity. After filtering, 3,616 P/LP variants were observed in NYC individuals. HGVS description and review status (gold stars) were obtained from variant_summary.txt.gz in https://ftp.ncbi.nlm.nih.gov/pub/clinvar last updated on March 30, 2024. The variants which were not classified as P/LP as of March 30, 2024 were removed from results.

We defined eight IBD groups with IBD scores >3 as founder populations. To identify founder variants, we set a conservative threshold, including only those that: a) were significantly enriched in a certain founder population compared with other NYC individuals (Fisher’s Exact p < 0.05), b) occurred at a MAF of <0.0001 in NYC individuals not assigned to that group, and c) appeared more than once in that group. We applied the Bonferroni correction (p value < 0.05 / (3,616 × 8)), but all results are listed in **Table 1** and **Table S2** since it is too strict for populations with small sample size. The minor allele frequencies of founder variants were extracted from gnomAD v3.1.225(47) using gnomAD_DB (https://github.com/KalinNonchev/gnomAD_DB) to compare frequencies in NYC dataset. The number of P/LP variants per individual was also counted for each IBD group.

### Ancestry analysis for the founder variant in the Caribbean IBD groups

We identified multiple IBD groups that appeared to have Caribbean ancestry, based on *F*_ST_ analysis against reference populations. Since Caribbean populations have three different continental ancestries in their genomes (African, Native American and European) due to their complex history, we inferred local ancestry around the founder variants detected in Caribbean populations shown in **Table 1** in order to reveal the ancestral background of those founder variants.

Genotype datasets for the carriers were generated by combining the genotype dataset used for IBD analysis and genotype data of each ClinVar variant, and phased by Beagle v5.4 without reference genomes. We then used RFMix(48) version 2 to infer local ancestry ±20 Mb of the variant, with 3 expectation-maximization steps. To assess local ancestry, we assembled a reference panel by identifying individuals from 1KG, HGDP and SGDP for whom >95% of their genomes appeared to have either African, American or European ancestry based on ADMIXTURE analysis with K=5 as reference (the same reference individuals in the SCOPE analysis).

## Data Availability

All statistic data produced in the present study are available upon reasonable request to the authors.

## ACKNOWLEDGEMENTS

The authors thank Drs. Alex Charney, Gillian Belbin, and Eimear Kenny for providing Bio*Me* self-described population ancestry labels. The advice of Humberto Brown (Director of Health Disparities, Arthur Ashe Institute for Urban Health) and of Karen Blanco and Katherine Oliva Blanco (Hondurans Against AIDS/Casa Yurumein) is also gratefully acknowledged. This work was supported by OT2OD031919 from the Office of the Director (NIH) to MS and SR and R01AG057422 from the National Institute on Aging (NIH) to JMG. We thank the All of Us Resource Access Board for their thoughtful review and final approval of this manuscript prior to publication. Finally, the authors thank the participants of AoU and Bio*Me*, without whom this research would not be possible.

The All of Us Research Program is supported by the National Institutes of Health, Office of the Director: Regional Medical Centers: 1 OT2 ODO26549; 1 OT2 ODO26554; 1 OT2 ODO26557; 1 OT2 ODO26556; 1 OT2 ODO26550; 1 OT2 OD26552; 1 OT2 ODO26553; 1 OT2 ODO26548; 1 OT2 ODO26548; 1 OT2 ODO2551; 1 OT2 ODO26555; IAA #: AOD 16037; Federally Qualified Health Centers: HHSN 263201600085U; Data and Research Center: 5 U2C ODO23196; Biobank: 1 U24 OD023121; The Participant Center: U24 ODO23176; Participant Technology Systems Center: 1 U24 ODO23163; Communications and Engagement: 3 OT2 ODO23205; 3 OT2 ODO23206; and Community Partners: 1 OT2 ODO25277; 3 OT2 ODO25315; 1 OT2 ODO25337; 1 OT2 ODO25276.

Molecular data for the Trans-Omics in Precision Medicine (TOPMed) program was supported by the National Heart, Lung and Blood Institute (NHLBI). Genome sequencing for “NHLBI TOPMed:phs001644 was performed at MGI (3UM1HG008853-01S2). Core support including centralized genomic read mapping and genotype calling, along with variant quality metrics and filtering were provided by the TOPMed Informatics Research Center (3R01HL-117626-02S1; contract HHSN268201800002I). Core support including phenotype harmonization, data management, sample-identity QC, and general program coordination were provided by the TOPMed Data Coordinating Center (R01HL-120393;U01HL-120393; contract HHSN268201800001I). We gratefully acknowledge the studies and participants who provided biological samples and data for TOPMed.

## Data availability

The data included in this manuscript was entirely obtained from publicly available resources. We have reported on our findings in accordance with the terms of the respective data sources (*e.g.* AoU requires that reporting on groups of individuals be restricted to ≥ 20 individuals). We have not generated data that requires sharing.

**Supporting information captions**

**Text S1: Supplementary methods**

**Figure S1: Ancestry background of AoU IBD clusters.**

**Figure S2: Comparison of NYC Census data in July 2022 and AoU NYC participants.**

**Figure S3: PCA plot for AoU NYC participants (a), BioMe (b) and global reference populations.**

**Figure S4: 16 IBD groups in the combined dataset of NYC.**

Included in TextS1_FigureSs.docx

**Table S1: ancestry background assignment to IBD groups**

**Table S2: All candidate founder P/LP variants in the eight founder populations in NYC**

**Table S3: Frequencies of Caribbean founder variants from Table 1 in other shared ancestry IBD groups**

Included in SupTable.xlsx

