## Supplemental text and figures for "Genetic disease risks of under-represented founder populations in New York City"

**Supporting Information**

**Table of contents**

**Supplementary methods**	2

**Subjects and dataset preparation** 	2

**Comparison of demography between US Census and AoU NYC participants**	2

**Population structure analysis for AoU**	2

PCA and global ancestry analysis for AoU	2

Identity-by-descent (IBD) analysis for AoU	3

Assessment of the founder effect	3

**IBD analysis for Bio*Me***	4

**Inferring ancestral background of individuals in IBD clusters** 	4

**Genetic ancestry analysis for the combined dataset** 	4

**Detection of pathogenic founder variants** 	5

**Ancestry analysis for the founder variant in the Caribbean IBD groups** 	5

**Supplementary figures**	6

**Figure S1**: Ancestry background of AoU IBD clusters.	6

**Figure S2**: Comparison of NYC Census data in July 2022 and AoU NYC participants.	7

**Figure S3**: PCA plot for AoU NYC participants (a), Bio*Me* (b) and global reference populations. 	8

**Figure S4:** 16 IBD groups in the combined dataset of NYC.	9

**References**	10

**Investigators**

Mariko Isshiki PhD,^1^ Anthony Griffen BSc,^2^ Paul Meissner MSPH,^3, 4^ Paulette Spencer MPH,^5^ Michael D. Cabana MD,^6^ Susan D. Klugman MD,^4^ Mirtha Colón MSW,^7^ Zoya Maksumova MD,^8^ Shakira Suglia ScD,^10^ Carmen Isasi MD,^6,9^ John M. Greally DMed,^1,6,†^ Srilakshmi M. Raj PhD^1,†^

**Text**

### Supplementary methods

#### 1. Subjects and dataset preparation

Array genotype data of the participants living in New York City (NYC) in the All of Us Program (AoU)^1^ version 6 curated data repository (CDR) were used to assess population structure of New York City participants using principal component analysis (PCA), global ancestry analysis by SCOPE^2^ and Identity-by-descent (IBD) analysis. The residence was defined based on 3-digit zip code provided by AoU. Samples were QCed by call rate and kinship coefficient using PLINK v2.00a2.3LM.^3^ No individuals were filtered out by the call rate threshold of 0.9 (---mind 0.1). Either of the pairs of individuals who showed king kinship coefficients > 0.125 were removed using --king-cutoff 0.125 in PLINK2. Variants of the array data were filtered with the following conditions using PLINK2: minor allele frequency >0.01, genotyping rate per site > 0.95, and p-value for the departures from Hardy Weinberg Equilibrium (HWE) > 1x10^-6^ (--maf 0.01 –geno 0.05 --snps-only --hwe 1e-06). After QC steps, 13,817 participants and 720,630 SNPs remained for downstream analysis.

Out of these individuals, 10,381 individuals had whole genome sequence data available. We used this subset of individuals and whole genome sequence data from an independent NYC biobank, the Mount Sinai Bio*Me* biobank,^4,5^ to identify founder pathogenic variants (see below).

2. **Comparison of demography between US Census and AoU NYC participants**

To show the extent to which our dataset represents the demography of NYC, we compared the proportion of four major self-described race and ethnicities per borough between census data and AoU NYC participants. We obtained census data from the following source:

<https://www.census.gov/quickfacts/fact/table/richmondcountynewyork,newyorkcountynewyork,queenscountynewyork,kingscountynewyork,bronxcountynewyork,newyorkcitynewyork/PST045222>.

For AoU NYC participants, self-identified race/ethnicity was obtained using a questionnaire. Participants answered the question: “Which categories describe you? Select all that apply. Note, you may select more than one group.” in the Basics Survey. Borough residence was defined based on the first three digits of the zip code of residence, provided by AoU.

3. **Population structure analysis for AoU**

To understand the population background of the AoU individuals, we used three approaches: 1) visualizing ancestry in relation to reference genomic data using principal components analysis (PCA), 2) global ancestry analysis to understand the ancestral background of each individual, and 3) identity-by-descent analysis to identify groups of individuals who appear to share ancestry.

We assembled a reference genomic dataset consisting of the 1000 Genomes project (1KGP),^6^ Simons Genome Diversity Project (SGDP),^7^ Human Genome Diversity Project (HGDP),^8^ and data from 10,000 individuals from the NYC-based Bio*Me* biobank^4,5^ collected through the Mount Sinai hospital system (dbGaP accession number: phs001644).

a. **PCA and global ancestry analysis for AoU**

PCA and global ancestry analysis were conducted on the dataset comprising 13,817 AoU participants and 3,584 individuals from 1KGP, HGDP and SGDP and 150,213 SNPs. To obtain the merged dataset, we first randomly sampled SNPs from the QCed AoU dataset comprising 13,817 participants and 720,630 SNPs. We then extracted those SNPs from the already QCed reference dataset which included all individuals except close relatives (filtered by KING kinship coefficient < 0.125) from 1KGP, HGDP and SGDP. The QC conditions of the reference dataset were the same as above: genotyping rate per person > 0.9, genotyping rate per site > 0.95, p-value for the departures from Hardy Weinberg Equilibrium (HWE) > 1x10^-6^ and biallelic SNPs. We merged the AoU dataset and reference dataset and removed close relatives again with KING kinship coefficient > 0.125.

PCA was performed with PLINK2.^3^ To measure global ancestry proportions, we ran SCOPE^2^ in supervised mode on the combined AoU and reference datasets.  We first conducted ADMIXTURE^9^ analysis with K=5 and identified individuals from 1KGP, HGDP and SGDP for whom >95% of their genomes appeared to have either of five continental ancestries as reference: African, European, South Asian, East Asian and Native American ancestries^6^. We then estimated allele frequencies for each reference super population with PLINK v1.90b6.22.^10^ We inferred global ancestry proportions corresponding to the reference populations using the PLINK output (the frq.strat output file) and the same genome-wide SNP dataset used for the PCA analysis.

b. **Identity-by-descent (IBD) analysis for AoU**

Identity-By-descent (IBD) segments are identical genomic regions shared between two individuals which were inherited from the common ancestors. IBD segments enabled us to identify disease associated variants,^5,11^ detect fine population structure,^4,12^ infer effective population size changes^13^ and examine the strength of a founder effect.^14^ As the IBD segments break down over generations due to recombination, it is a powerful tool to capture demographic movement in the recent past. In a founder population which has experienced small population size in its recent past, individuals are more likely to share recent common ancestors; that is, they are likely to share more IBD segments.

We phased the QCed array data comprising 13,817 participants and 720,630 SNPs with Beagle v5.4^15^ using all populations from 1KGP phase 3 dataset^6^ as reference. TPBWT^16^ was performed on the phased dataset to infer IBD segments >3 cM across all pairs of individuals using the default settings. This segment length threshold was set to obtain reliable estimation of IBD tracts through TPBWT, as the accuracy of TPBWT was lower in shorter length IBD segments especially when inferred using whole genome sequences.^17^ The same threshold was set when estimating IBD segments in the whole genome sequence data from Mount Sinai Bio*Me* Biobank participants (see below).

We identified 39,105,629 shared IBD segments for 17,821,529 pairs of individuals. The mean amount of IBD sharing per pair was 12.15 cM. The length of IBD segments (cM) shared between each pair of individuals were summed to obtain the total length of IBD sharing between pairs. We used this cumulative IBD sharing for all pairs in our dataset to construct an undirected network using the iGraph package^18^ in R. We found several pairs of individuals with shared IBD segments > 500cM, which placed their mean IBD sharing value in the 75% quantile or above, when compared to all other pairs of individuals in the dataset. These individuals could be relatives who were not excluded by the kinship coefficient threshold. Therefore, in order to focus on recent demography and to reduce clustering of extended families, we filtered for edges with cumulative IBD sharing ≥12 cM and ≤72 cM, as previously described.^19,20^ Community detection was performed using the infomap.community() function^21^ from the iGraph package on the constructed undirected network where each node represented an individual, and edge weights were defined as the total IBD sharing between individuals. We chose the infomap algorithm following the approach of Belbin et al. (2021),^4^ in which they identified population substructure in Bio*Me*. Every individual was assigned to only one cluster. We detected 386 IBD clusters in total. The number of individuals belonging to each cluster varied substantially, ranging from 1 individual to 3715 individuals. We detected 14 clusters containing 20 and more individuals, which covered 91% of the AoU participants, and used these clusters for downstream analyses.

c. **Assessment of the founder effect**

To test the strength of founder effect for each IBD cluster, we estimated the ‘IBD score’, the average length of IBD segments between 3–20 centimorgans (cM) shared between two genomes normalized to sample size, as previously described.^14^ We also estimated the ‘IBD score’ per cluster per borough.

4. **IBD analysis for Bio*Me***

To confirm the robustness of our approach and to obtain good references for population annotation, we conducted IBD sharing network analysis for an independent NYC cohort, the Mount Sinai Bio*Me* Biobank. The original authors also conducted IBD analysis on their full cohort (n= 32,595 ) in Belbin et al. (2021),^4^ and they identified 17 clusters that each contained at least 100 individuals. We obtained whole genome sequence data of 12,022 Bio*Me* participants from dbGaP (accession no. phs001644), which was sequenced in Trans-Omics for Precision Medicine (TOPMed) Freeze 9. Variant QC was then conducted using VCFtools v0.1.13^22^ with the following conditions: minor allele frequency >0.05, genotyping rate per site > 0.95, p-value for the departures from Hardy Weinberg Equilibrium (HWE) > 1e-6, and after passing all these filters, only including biallelic SNPs (---maf 0.01 --max-missing 0.95 -hwe 0.000001 --remove-filtered-all --remove-indels --min-alleles 2 --max-alleles 2). To reduce the computational burden, we randomly selected one-quarter of the SNPs (approximately 1,000,000 SNPs for the whole genome). Phasing was conducted with Beagle v5.4^15^ with the combined reference genome dataset as described above. Either of the pairs of individuals who showed KING kinship coefficients > 0.125 were removed using PLINK2 --king-cutoff 0.125. The final dataset consisted of 11,549 individuals and 982,770 SNPs. We estimated IBD segments, constructed an IBD sharing network and estimated IBD scores as described above. We identified 39,598,121 shared IBD segments for 14,988,829 pairs in total. The mean amount of IBD sharing per pair was 14.59 cM, which was similar to the amount of IBD sharing in our AoU dataset. We identified 16 clusters with 20 and more individuals, which covers 92.5% of BioMe participants. We annotated each cluster based on self-identified ancestry information kindly provided by Bio*Me* leadership (Alexander Charney, personal communication) (**Table S1**).

5. **Inferring ancestral background of individuals in IBD clusters**

The AoU research program does not provide information about the country of origin of participants to AoU workbench users at this stage. To infer the genetic ancestry of individuals in our defined IBD clusters, we estimated Hudson’s *F*_ST_ between each cluster and populations in reference panels using PLINK2. We used a reference panel combining the reference panel used for PCA and global ancestry analysis and QCed genotype data of Bio*Me* individuals, who all had self-described population ancestry labels. Close relatives were removed after merging AoU dataset and the merged reference panel using PLINK2 -–king-threshold 0.125. The final dataset included 28,802 individuals and 140,952 biallelic SNPs. To obtain robust results, F_ST_ values were inferred with populations with ≥ 10 individuals. A heatmap was drawn with Python Matplotlib^23^ and Seaborn^23^ packages implemented in Python 3.8.8.

6. **Genetic ancestry analysis for the combined dataset**

We also conducted PCA analysis for the merged dataset of AoU and Bio*Me*. We combined IBD clusters in AoU and Bio*Me* where the *F*_ST_ value between a pair of IBD clusters was < 0.001, resulting in 16 clusters in NYC. *F*_ST_ values between the combined clusters were estimated to see the relationship between clusters and used to write network of IBD populations. The network was plotted with igraph package implemented in R using log_10_(*F*_ST_) as a distance matrix. Only edges with *F*_ST_ <1x10^-2^ were shown in **Figure 2**.

7. **Detection of pathogenic founder variants**

As of Jan 7, 2023, 1,577,987 variants were registered in ClinVar. Of these, we identified 193,935 pathogenic variants whose CLINSIG flags started with “Pathogenic” or “Likely Pathogenic”. We extracted these variants from whole genome sequence data of 10,381 NYC All of Us participants using Hail version 0.2.107-2387bb00ceee^24^ and from those of BioMe participants using vcftools version 0.1.13. We merged the dataset using PLINK v1.90b6.22 --bmerge, removed close relatives with KING kinship coefficient > 0.125 with PLINK2 –king-threshold 0.125 and excluded variants which appeared less than twice in our combined dataset with PLINK2 flag --mac 2. Variants with genotype rate < 0.9 and p-value for the departures from Hardy Weinberg equilibrium <1e-16 were filtered out with PLINK2. We set a small Hardy Weinberg equilibrium threshold since rare variants are likely to diverge from Hardy Weinberg equilibrium due to high heterozygosity in a general population sample like the whole dataset of AoU and Bio*Me*. After filtering, 3,616 pathogenic variants were observed in NYC individuals’ whole genome sequence data.

We defined eight IBD clusters with IBD score >3 as founder populations. The frequency of the variants in each founder IBD cluster was estimated using PLINK v1.9. We set a conservative threshold to identify founder variants, including only those that were: a) Significantly enriched in a certain founder population against other NYC individuals after Bonferroni correction (Fisher’s Exact p value < 0.05), b) minor allele frequency (MAF) <0.0001 in NYC individuals not assigned to that cluster and c) appeared more than once in that cluster. We applied the Bonferroni correction (p value < 0.05 / (3,616 x 8)), but all results are listed in **Table 1** and **Table S2** since it is too strict for populations with small sample size. We checked and confirmed the relatives in a closer relationship than our threshold (KING kinship coefficient = 0.125) did not affect our results by applying an additional --king-threshold 0.0884.

The number of pathogenic variants per individual is also counted for each IBD cluster to examine if there are differences in the distribution of known pathogenic variants based on ancestral background.

The minor allele frequencies of founder variants were extracted from gnomAD v3.1.2^25^ using gnomAD_DB(https://github.com/KalinNonchev/gnomAD_DB) to compare frequencies in NYC dataset.

8. **Ancestry analysis for the founder variant in the Caribbean IBD groups**

We identified multiple IBD clusters that appeared to have Caribbean ancestry, based on *F*_ST_ analysis against reference populations. Since Caribbean populations have three different continental ancestries in their genomes (African, Native American and European) due to their complex history, we inferred local ancestry around the founder variants detected in Caribbean populations shown in **Table 1** in order to reveal the ancestral background of those founder variants.

Genotype datasets for the carriers were generated by combining the genotype dataset used for IBD analysis and genotype data of each ClinVar variant, and phased by Beagle v5.4 without reference genomes since most of the variants were not found in the reference. When the number of the ClinVar variant carriers were less than 50 individuals, 50 individuals from that founder population were analyzed together to maintain accuracy for phasing. We then used RFMix^26^ version 2 to infer local ancestry ±20 Mb of the variant, with 3 expectation-maximization steps. To assess local ancestry, we assembled a reference panel by identifying individuals from 1KG, HGDP and SGDP for whom >95% of their genomes appeared to have either African, American or European ancestry based on ADMIXTURE analysis with K=5 as reference (the same reference individuals in the SCOPE analysis).

### Figures

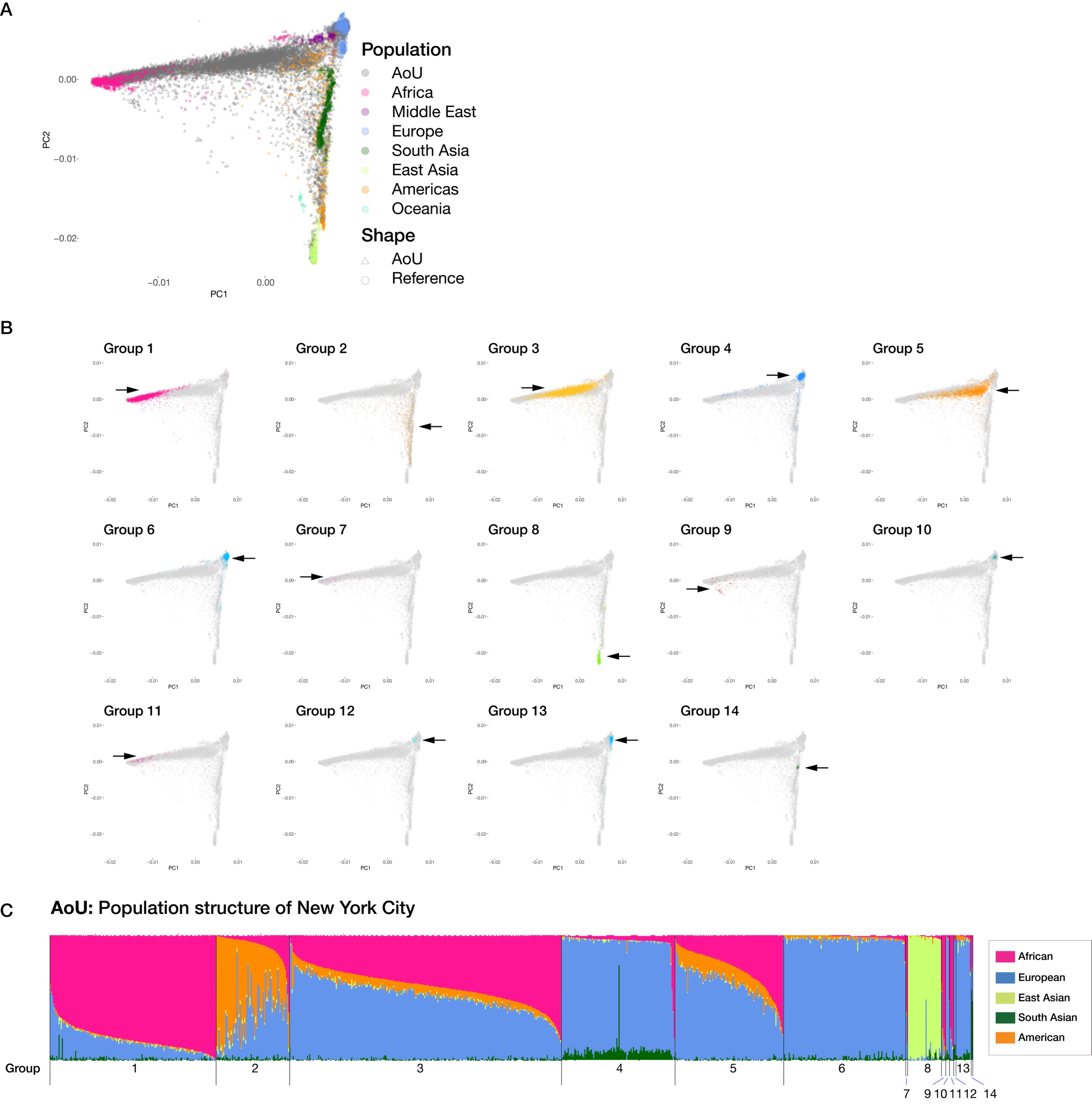

**Figure S1**

Ancestry background of AoU IBD clusters.

**(a)** PCA plot for all AoU NYC participants and reference panels. **(b)** PCA plots highlighting individuals belonging to the 14 IBD clusters detected in AoU NYC participants. **(c)** SCOPE analysis for AoU NYC participants labelled with the 14 IBD clusters. Each color represents global ancestry proportion of the five superpopulations (African, European, American, East Asian and South Asian) inferred using supervised mode in SCOPE.

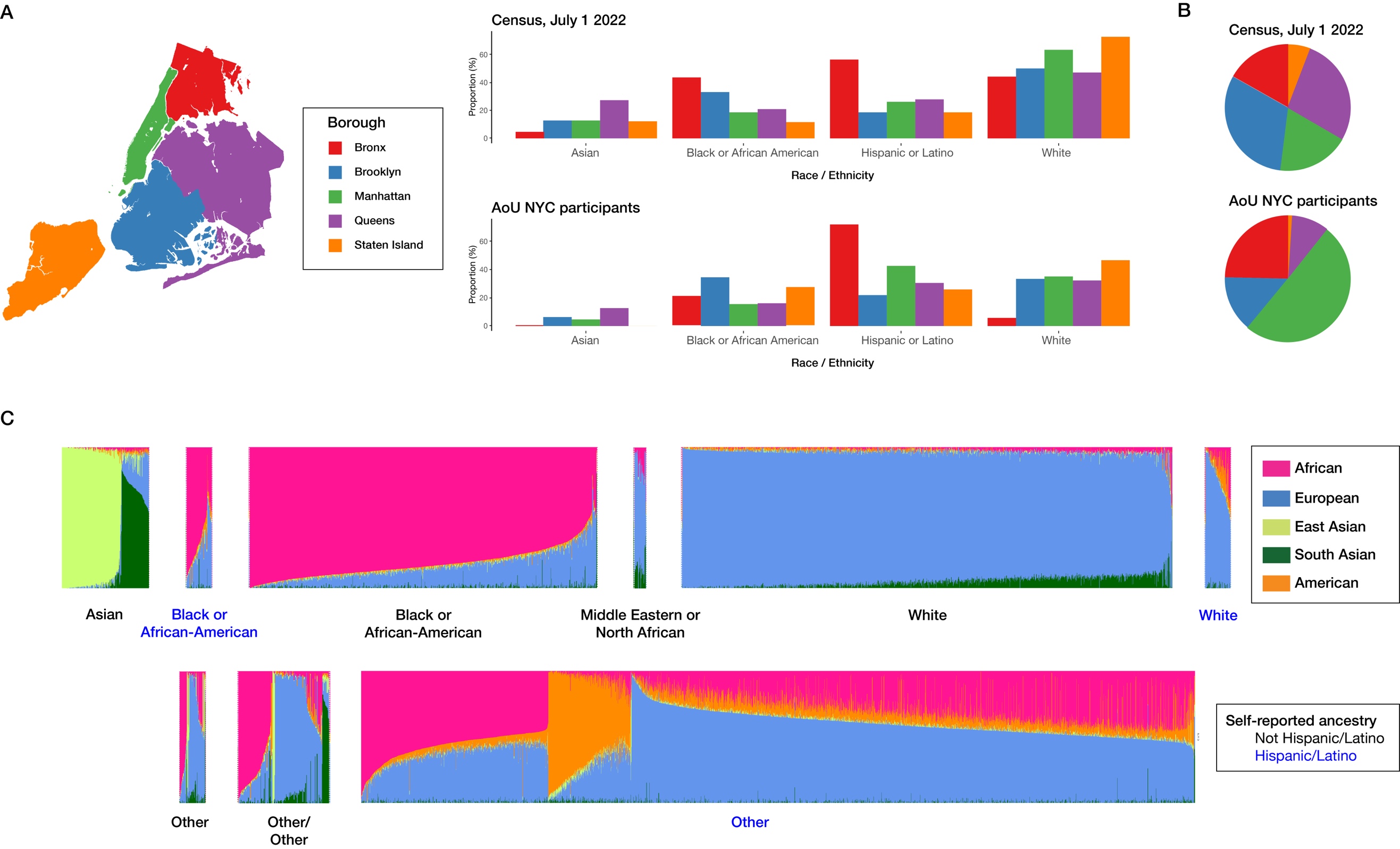

**Figure S2**

Comparison of NYC Census data in July 2022 and AoU NYC participants

**(a)** The proportion of four major races and ethnicities (Asian, Black or African American, Hispanic or Latino and White) in each borough according to the US census (July 2022) and AoU NYC participants. **(b)** Proportional US census population sizes compared with the proportions of AoU participants from each borough. Although the AoU dataset had disproportionately higher Manhattan and Bronx participants, it showed similar proportions of the four race and ethnicity categories in each borough compared with census data.

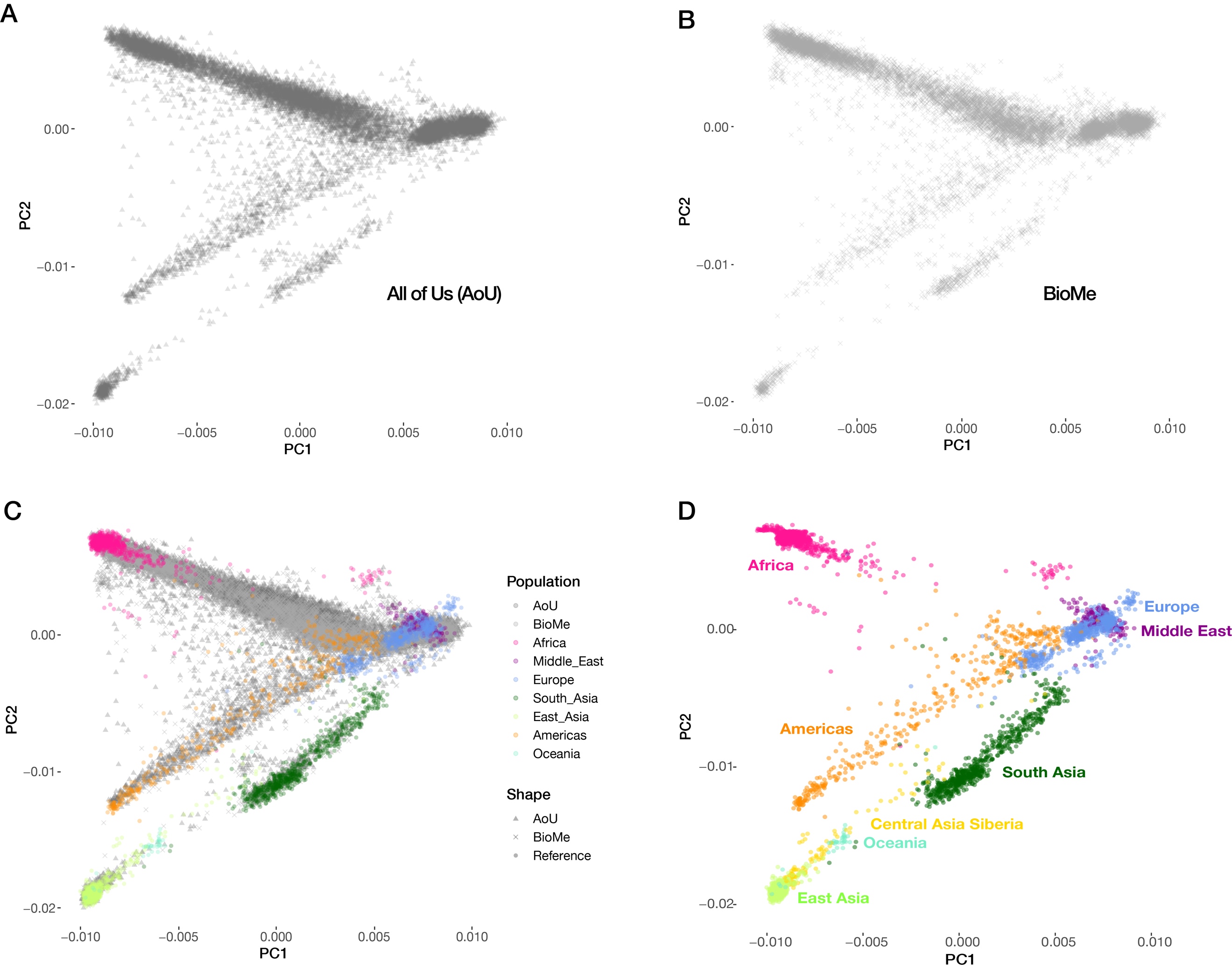

**Figure S3**

PCA plot for **(a)** AoU NYC participants, **(b)** Bio*Me* participants, **(c)** both cohorts compared with global reference populations, and **(d)** the global reference populations alone.

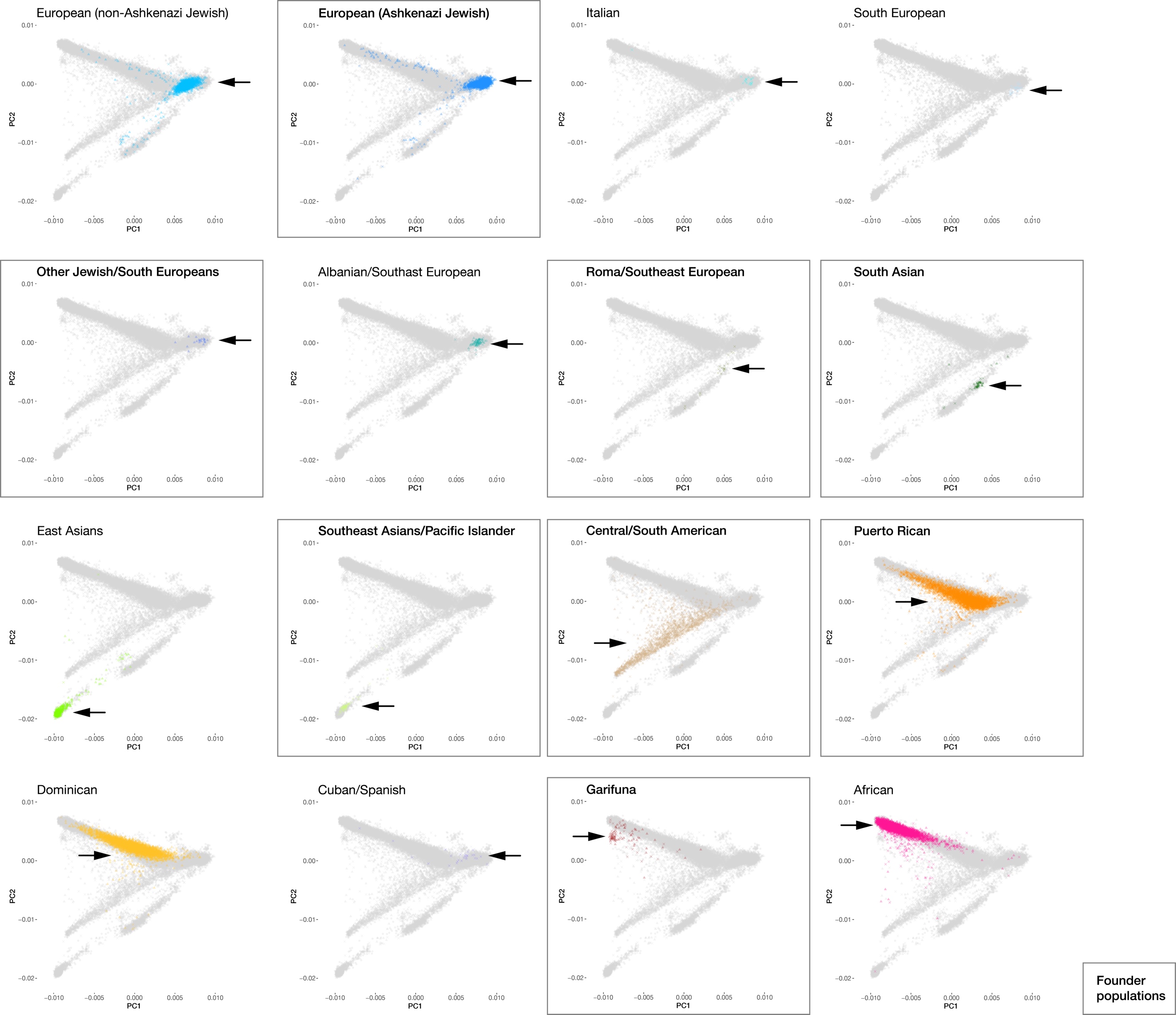

**Figure S4**

PCA plots for the 16 IBD clusters from **Figure 1c**, with the 8 founder populations highlighted in boxes.
